# The unexplainable plasma protein measurements from Proximity Extension Assays

**DOI:** 10.64898/2026.08.10.26360077

**Authors:** Justus F. Gräf, Nigel Kurgan, Simon Rasmussen

## Abstract

Plasma proteomics assays aim to capture biologically interpretable circulating protein signals. Here we systematically assess Olink Explore targets with exceptionally low variance explainability by integrating genetic associations, cross-platform concordance and tissue and peptide atlases. We identify a subset of protein targets likely lacking robust plasma signals, highlighting assay- and tissue-specific limitations with implications for panel design, statistical power and interpretation of large-scale proteomics studies.

## Main

Proximity Extension Assays, including the Olink technology, are now widely used in large population proteomics studies, yet thorough technical validation of measured targets remains limited. Although many of the targets show strong biological and clinical associations, for example in the UK Biobank (Pietzner *et al*, 2025a; Sun *et al*, 2023; Sigurdsson & Gräf *et al*, 2025), the reliability and interpretability remain a topic of debate in the field. This is largely caused by reports on discordant associations with other proteomic platforms, like mass spectrometry (MS)-based proteomics or aptamer-based platforms like SomaScan (Carrasco Zanini *et al*, 2025; Eldjarn *et al*, 2023; Kirsher *et al*, 2025).

A recent study by Pietzner et al., (Pietzner *et al*, 2025a) systematically deconvolutes determinants of plasma protein levels measured using the Olink Explore platform, providing valuable insight into the variance and explainability of these protein measurements. Their work demonstrated that most protein targets show substantial biological signals indicated by their strong association with a wide range of clinical and molecular traits, significantly improving interpretability of the measured plasma proteins in large cohorts like the UK Biobank (Sun *et al*, 2023). However, this analysis also revealed a subset of proteins with unexpectedly low variance explained (<1%) by biological or technical factors (Pietzner *et al*, 2025a). While this could be due to limited availability of features in the UK Biobank, it might also raise questions about assay measurement reliability and interpretability. To examine this, we integrated published data on proteomic platform comparisons, genetic associations, and tissue and peptide atlases to assess whether these measurements represent meaningful biological signals despite low explainability or potentially harbor assay artefacts.

We identified 411 proteins in the Olink Explore panel with less than 1% of total variance explained in the Pietzner et al. (Pietzner *et al*, 2025a) dataset. Such minimal variance is atypical for molecular traits that are often influenced by genetic, clinical, biological, or technical parameters. Of these proteins, 283 (69%) do not show any detectable cis- or trans-pQTLs in the largest published genetic studies of plasma proteins measured by this assay (**Figure 1a**) (Sun *et al*, 2023; Eldjarn *et al*, 2023; Hawkes *et al*, 2025; Dhindsa *et al*, 2023; Cai *et al*, 2025). In total, 1,838 targets from the Olink Explore panel used in the UK Biobank were also measured in an Icelandic cohort and compared to SomaScan measurement of the same targets, which includes 115 of the 283 proteins with <1% variance explained and no pQTLs. Strikingly, most of these 115 proteins show near-zero correlation across the two affinity-based platforms (Eldjarn *et al*, 2023), in contrast to the strong cross-assay concordance observed for many proteins (**Figure 1b**). While neither low-explainability, lack of genetic association nor low correlation between assays alone necessarily indicate that these measurements are inaccurate, the combination of the datasets suggest that a subset of up to 152 targets (5.2%) from the Olink Explore panel may not currently provide biologically interpretable or quantitative information with evidence from all three datasets for 96 targets (3.3%) (**Figure 1c, Supplementary Table**).

**Figure 1.**
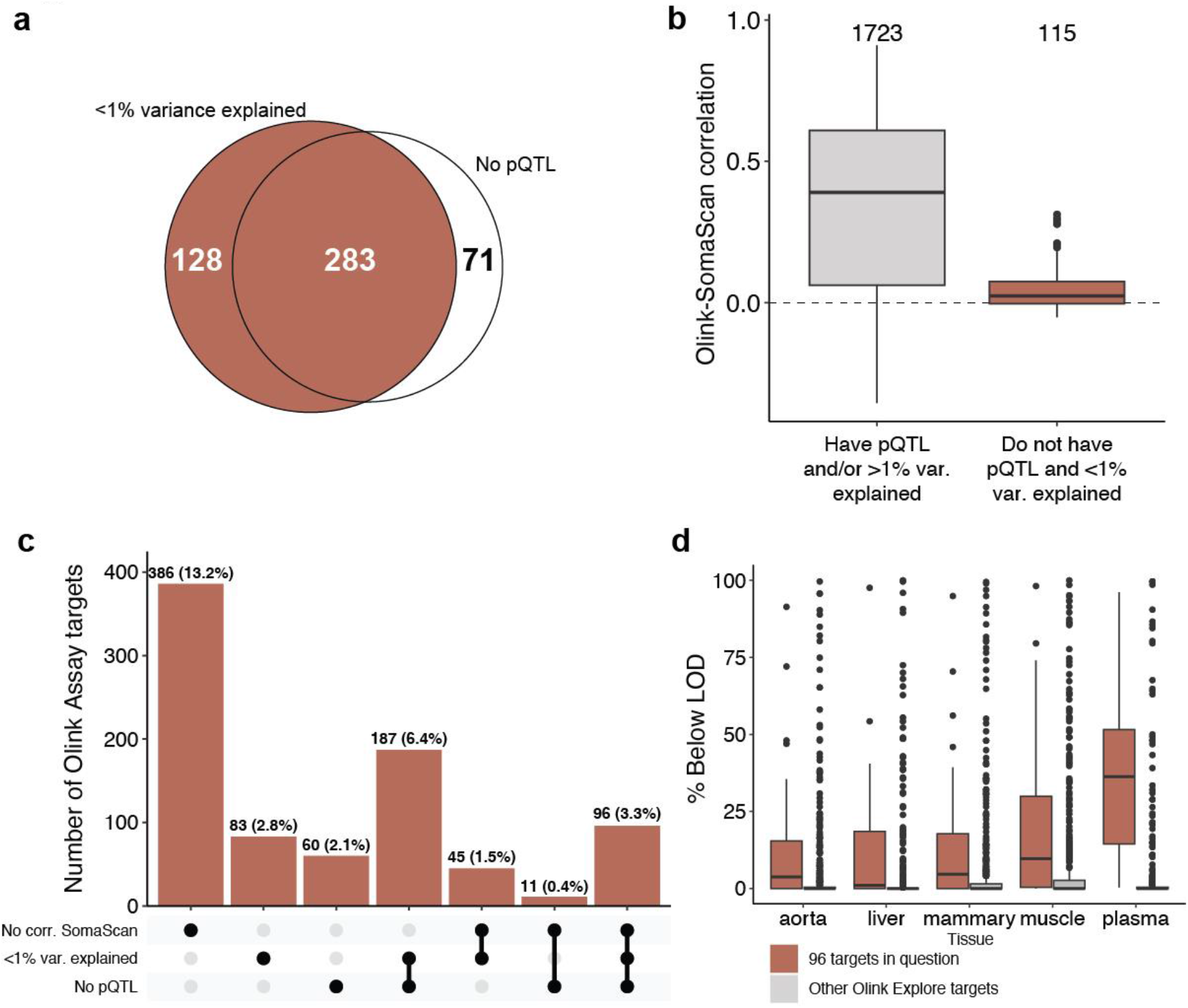
**a)** Venn diagram illustrating the number of Olink Explore targets with <1% variance explained from Pietzner et al., (n=488) overlapped with Olink Explore targets without pQTL (n=354) from Sun et al. 2023, Dhindsa et al. 2023, Hawkes et al. 2025 and Cai et al. 2025. **b)** Boxplot showing Spearman correlation between Olink Explore and SomaScan measurements for the Olink Explore targets that are overlapping with SomaScan measurements in Eldjarn et al., 2023 (n=1,838 / 2,922) stratified by if they have pQTL and >1% variance explained in Pietzner et al. 2025. **c)** Number of Olink Explore targets in different intersection sets dependent on correlation with SomaScan (<0.1 Spearman correlation), explained variance and pQTL. The percentage indicates the fraction of targets compared to all Olink Explore targets (n=2,922). **d)** Percentage of Olink Explore assay targets falling below limit of detection (LOD) measured in different tissues by Hartley et al., 2025, stratified by if the targets are amongst the 96 targets with low-explainability identified here.

To investigate why certain Olink-measured proteins exhibit low explainability, we evaluated three factors: (1) how confidently these 96 proteins are detected in blood, (2) whether they show tissue-specific enrichment, and (3) whether they possess pQTLs identified by MS-based proteomics or SomaScan. The Human Peptide Atlas (Desiere *et al*, 2006) provides robust MS-based evidence for proteins confidently detected in plasma. Among the 96 low-explainability proteins, evidence for plasma detection was markedly depleted (23% [23/96] vs. 54% [1522/2827], OR = 0.27, p = 5.88×10^−8^). Utilizing the Genotype-Tissue Expression (GTEx) atlas, we also found that these proteins were enriched for brain-derived (13% [12/96] vs. 4% [100/2827], OR = 3.9, q = 2.3×10^−4^) and reproductive-organ enriched proteins (7% [7/96] vs. 2% [50/2827], OR = 3.96, p = 0.035), suggesting limited biological relevance within the circulation. Consistently, MS-based pQTLs were less common among these proteins compared to the comparison set (1% [1/96] vs. 17% [467/2827], OR = 0.053, p = 1.4×10^−6^) (Niu *et al*, 2023; Suhre *et al*, 2024; Pietzner *et al*, 2025b; Xu *et al*, 2023; Suhre *et al*, 2025). However, SomaScan data from the Icelandic deCODE cohort (35,559 individuals) revealed pQTLs for 97% (93/96) of these proteins, compared with 60% (1681/2827) in the remaining proteins (OR = 21.1, p = 3.98×10^−17^) (Eldjarn *et al*, 2023; Ferkingstad *et al*, 2021). This suggests that SomaScan captures biologically meaningful signals for these proteins that neither Olink nor MS-based proteomics can readily detect.

One explanation for this large discrepancy in detection of biological signal for these selected proteins is that they are more conducive to the small footprint of aptamer (SomaScan) binding compared to large antibodies (Olink). This hypothesis is supported by recent multi-tissue Olink data (Hartley *et al*, 2025) which found plasma-specific variability in protein detection of these 96 proteins relative to tissue detection (**Figure 1d**). These platform and tissue-specific findings may partially be explained by a higher proportion of interleukins (7% [7/96] vs 1% [23/2827], OR = 9.59, p = 7.98×10^−4^) found within this subset of proteins or immunoglobulin interference of antibody binding affinity in plasma (Wik *et al*, 2021).

Collectively, these findings raise concerns about the reliability of Olink plasma measurements for this subset of proteins. Including such targets in high-cost panels may offer limited biological value and increase type II error if their circulating levels cannot be robustly quantified. Importantly, the ability of SomaScan to detect pQTLs does not necessarily confirm it is measuring the intended analytes; rather, it underscores discrepancies between affinity-based platforms. Resolving what Olink and SomaScan are each capturing and why proteins with weak or biologically implausible plasma detectability can reach genome-wide significance in one platform but not the other, requires orthogonal validation. Approaches such as immunoprecipitation coupled to MS, targeted MS assays, direct assessment of capture reagent specificity (Olink antibodies and SomaScan aptamers), and deeper characterization of tissue contribution to the plasma proteome in health and disease (Malmström *et al*, 2025) will be essential for establishing the analytical validity and value of these measurements.

## Online Methods

### Identification of the low-explainability Olink plasma proteome

We identified 411 proteins in the Olink Explore panel with <1% of total variance explained in the Pietzner et al. (2025a) dataset. These proteins were overlapped with the largest published genetic studies of Olink Explore plasma proteins to determine pQTL status, yielding 283/411 with no detectable cis- or trans-pQTLs (Sun et al., 2023; Eldjarn et al., 2023; Hawkes et al., 2025; Dhindsa et al., 2023; Cai et al., 2025). We then intersected this set with an Icelandic cohort comparing SomaScan and Olink measurements (Eldjarn et al., 2023); 115/283 had overlapping SomaScan data. The “core” unexplainable Olink plasma proteins were defined as those with no variance explained, no pQTL, and no significant SomaScan-Olink correlation (n = 96; olink_assay_summary_evidence_inaccuracies.xlsx), and all remaining assays were treated as the “other” comparison set.

### Integration of tissue, protein class, and other pQTL evidence

Evidence sources were mapped to the “core” and “other” protein lists by UniProt ID, with gene-symbol used when UniProt IDs were unavailable. Evidence for detection in plasma was defined using the human plasma PeptideAtlas release 2025.08 (Desiere, 2006). Tissue enrichment evidence was obtained from the [GTEx Portal](https://www.gtexportal.org/home/) on 12/01/2025 (gtex_tissue_enrichment.xlsx), extracting how many “core” and “other” proteins were enriched for each tissue. Plasma pQTL evidence was compiled from large MS-based studies (Niu et al., 2023; Suhre et al., 2024; Pietzner et al., 2025b; Xu et al., 2023; Suhre et al., 2025) and SomaScan studies (Eldjarn et al., 2023; Fang et al., 2025). Tissue-specific limit-of-detection (% below LOD) and tissue pQTL annotations for Olink were extracted from Hartley et al. Human Protein Atlas v24 (Uhlén et al., 2015) was used for protein class annotations (e.g., interleukin enrichment).

### Statistics

For each binary annotation, we computed proportions in core versus other proteins and performed Fisher’s exact tests to estimate odds ratios and two-sided p-values; continuous features (e.g., % below LOD, number of GTEx tissues) were summarized by group means. We applied Benjamini-Hochberg correction separately within each set of tests, including GTEx tissue-specific enrichment and HPA protein class enrichment.

## Data Availability

A Supplementary Table with Olink Explore targets annotated with the summarized findings from this study is provided.

https://github.com/RasmussenLab/unexplainable_plasma_proteome).

## Data and code availability

All scripts and published data used in this study can be accessed on Github (https://github.com/RasmussenLab/unexplainable_plasma_proteome). A Supplementary Table with Olink Explore targets annotated with the summarized findings from this study is provided.

## Acknowledgements

S.R., J.F.G., and N.K. were supported by the Novo Nordisk Foundation (NNF23SA0084103). J.F.G. was supported by a research grant from the Danish Cardiovascular Academy, which is funded by the Novo Nordisk Foundation, grant number NNF20SA0067242 and The Danish Heart Foundation. N.K. is supported by the BRIDGE – Translational Excellence Programme (bridge.ku.dk) at the Faculty of Health and Medical Sciences, University of Copenhagen, funded by the Novo Nordisk Foundation (Grant agreement no. NNF23SA0087869).

## Author contributions

Conceptualization: J.F.G., N.K. Formal Analysis: J.F.G., N.K. Investigation: J.F.G., N.K. Methodology: J.F.G., N.K. Resources: S.R. Software: J.F.G., N.K. Supervision: S.R. Visualization: J.F.G. Writing – original draft: J.F.G., N.K. Writing – review & editing: J.F.G., N.K., S.R.

